# Topologic Parametric Response Mapping Identifies Tissue Subtypes Associated with Emphysema Progression

**DOI:** 10.1101/2023.06.16.23291508

**Authors:** Jennifer M. Wang, Alexander J. Bell, Sundaresh Ram, Wassim W. Labaki, Benjamin A. Hoff, Susan Murray, Ella Kazerooni, Stefanie Galban, Charles R. Hatt, MeiLan K. Han, Craig J. Galban

## Abstract

**Rationale and Objectives:** Small airways disease (SAD) and emphysema are significant components of COPD, a heterogenous disease where predicting progression is difficult. SAD, a principal cause of airflow obstruction in mild COPD, has been identified as a precursor to emphysema. Parametric Response Mapping (PRM) of chest computed tomography (CT) can help distinguish SAD from emphysema. Specifically, topologic PRM can define local patterns of both diseases to characterize how and in whom COPD progresses. We aimed to determine if distribution of CT-based PRM of functional SAD (fSAD) is associated with emphysema progression.

**Materials and Methods:** We analyzed paired inspiratory-expiratory chest CT scans at baseline and 5-year follow up in 1495 COPDGene subjects using topological analyses of PRM classifications. By spatially aligning temporal scans, we mapped local emphysema at year 5 to baseline lobar PRM-derived topological readouts. K-means clustering was applied to all observations. Subjects were subtyped based on predominant PRM cluster assignments and assessed using non-parametric statistical tests to determine differences in PRM values, pulmonary function metrics and clinical measures.

**Results:** We identified distinct lobar imaging patterns and classified subjects into three radiologic subtypes: emphysema-dominant (ED), fSAD-dominant (FD), and fSAD-transition (FT: transition from healthy lung to fSAD). Relative to year 5 emphysema, FT showed rapid local emphysema progression (−57.5% ± 1.1) compared to FD (−49.9% ± 0.5) and ED (−33.1% ± 0.4). FT consisted primarily of at-risk subjects (roughly 60%) with normal spirometry.

**Conclusion:** The FT subtype of COPD may allow earlier identification of individuals without spirometrically-defined COPD at-risk for developing emphysema.

## INTRODUCTION

Emphysema, defined as alveolar destruction and airspace enlargement distal to the terminal bronchiole, is a characteristic pathologic process of COPD [1]. Numerous studies using computed tomography (CT) for quantifying emphysema have reported the association of emphysema with a decline in lung function [2, 3] and functional status, increased dyspnea [4, 5] and overall worse clinical outcomes [6, 7]. The significance of emphysema on patient health is evident in a study by Zulueta and colleagues [7], who found in a large cohort of asymptomatic smokers that emphysema, quantified using a scoring method on CT, predicts early death from COPD and lung cancer. This finding highlights the importance of early diagnosis of emphysema. Although quantitative emphysema detection methods exist [8-12], these techniques only identify the presence of emphysema and not its onset.

Small airways disease (SAD), another major contributor to pulmonary obstruction in COPD, has been identified as a potential precursor to emphysema. Using microCT, McDonough and colleagues identified narrowing and destruction of smaller airways along the periphery of emphysematous regions of lung tissue. Based on these observations, they suggested that SAD is an early lesion that develops prior to emphysema [13]. Similar conclusions were proposed by Galban et al. using Parametric Response Mapping (PRM), a paired inspiratory and expiratory CT technique that indirectly measures SAD even in the presence of emphysema [14]. It has been previously demonstrated that PRM-derived SAD, referred to as functional small airways disease (PRM^fSAD^), is an independent predictor of lung function decline and that regions of PRM^fSAD^ do transition to emphysema [14, 15].

Due to the delay of symptoms in emphysema and SAD, these diseases are diagnosed late. As such, little is known about the local progression patterns in emphysema. In this study, we investigated how PRM-based readouts can identify areas of local lung parenchyma with progressive emphysema. Our quantitative CT method is an extension of our PRM approach, which can provide detailed local information on the distribution and arrangement of PRM-derived fSAD and normal parenchyma to identify distinct radiologic patterns associated with emphysema progression.

## MATERIALS AND METHODS

### Study Sample

Our study was a secondary analysis of data from COPDGene (ClinicalTrials.gov: NCT00608764), a large NIH-funded prospective multi-center observational study. In Phase 1 (2007-2012) of the original study, written and informed consent was obtained from all participants and the study was approved by local institutional review boards of all 21 centers. Ever-smokers with greater than or equal to 10 pack-year smoking history, with and without airflow obstruction, were enrolled between January 2008 and June 2011. For Phase 2 (2012-2017), participants were invited to return for a follow up evaluation. Approximately half of the Phase 1 cohort returned for the 5-year follow up visit (Phase 2). Participants were non-Hispanic white or African American. Participants underwent volumetric inspiratory and expiratory CT using standardized protocol; images were transferred to a central lab for protocol verification and quality control [16]. Exclusion criteria for COPDGene included a history of other lung disease (except asthma), prior surgical excision involving a lung lobe or greater, present cancer, metal in the chest, or history of chest radiation therapy. Participants were excluded from the present study due to inadequate CT for computing topologic parametric response mapping (tPRM), such as missing an inspiration/expiration scan, or failing quality control implemented specifically for the present study. Quality control protocol is described in **Supplemental Figure 1**. Data for participants evaluated here have been utilized in numerous previous studies, and a list of COPDGene publications can be found at http://www.copdgene.org/publications.htm. Our study is the first to report on tPRM analysis across the entire Phase 1 and Phase 2 cohorts of COPDGene participants.

### Subject Characteristics, Spirometry and CT Imaging

Subject characteristics, spirometry and CT imaging were acquired from all subjects at Phase 1 and 2. Spirometry was performed in the COPDGene study before and after the administration of a bronchodilator, specifically 180 mcg of albuterol (Easy-One spirometer; NDD, Andover, MA). Post-bronchodilator values were used in our analyses. COPD was defined by a post-bronchodilator FEV_1_/FVC of less than 0.7 at the baseline visit, as specified in the GOLD guidelines [17]. GOLD grades 1-4 were used to define spirometric disease severity. GOLD 0 classification was defined by a post-bronchodilator FEV_1_/FVC ≥ 0.7 at the baseline visit, alongside FEV_1_% predicted ≥ 80%. Participants with FEV_1_/FVC ≥ 0.7 with FEV_1_% predicted less than 80% were classified as having preserved ratio impaired spirometry (PRISm) [18]. In addition, demographics and smoking history were collected and 6-minute walking distance was measured. Health-related quality of life was assessed via St. George’s Respiratory Questionnaire (SGRQ) [19]. All CT data were obtained and analysis was performed as part of the COPDGene project. Whole-lung volumetric multidetector CT acquisition was performed at full inspiration (total lung capacity) and normal expiration (functional residual capacity) using a standardized previously published protocol [16]. Data reconstructed with the standard reconstruction kernel was used for quantitative analysis and all CT data were presented in Hounsfield units (HU) [16].

### Parametric Response Map (PRM)

Parametric Response Mapping was performed on all paired CT scans using Lung Density Analysis (LDA) software (Imbio, LLC, Minneapolis, MN) to generate PRM maps. In brief, LDA segmented the lungs and lobes with airways removed and inspiratory CT scans were spatially aligned to the expiratory images. Lung voxels were classified using pre-determined HU thresholds as: normal (PRM^Norm^, -950 < inspiration HU ≤ -810, and expiration HU ≥ -856), functional small airways disease (PRM^fSAD^, -950 < inspiration HU ≤ -810, expiration HU < -856), emphysema (PRM^Emph^, inspiration HU < -950, expiration HU < -856), or parenchymal disease (PRM^PD^, inspiration HU > -810) [20]. Only voxels between -1,000 HU and -250 HU at both inspiration and expiration were used for PRM classification.

### Topology Analysis of PRM

Topological analysis of PRM (tPRM) was performed using methods previously described [21]. tPRM metrics were defined through application of Minkowski measures on 3D binary voxel distributions: volume density (V), surface area (S), mean breadth (B), and Euler-Poincaré characteristic (𝜒) [22]. Maps of Minkowski measures (V, S, B, 𝜒) were computed for each PRM class map. Summary tPRM values for each participant were computed as the mean tPRM value of voxels over the entire lung volume. To indicate the PRM class associated with a Minkowski measure, the PRM class is presented as a superscript (e.g., V^fSAD^ is the volume density of PRM^fSAD^). tPRM analysis was performed using open-source and in-house software developed in MATLAB R2019a (MATLAB, The MathWorks Inc., Natick, MA).

### Alignment of Regional Emphysema at Year 5 to Baseline tPRM

To evaluate baseline tPRM values within regional emphysema defined at year 5, the following process was performed: 1. Volume density maps of PRM^Emph^ were determined using year 5 paired CT scans (V^Emph5^); 2. Volumes of interest were defined as having a V^Emph5^ > 0.25, a volume at year 5 equivalent to a 10 mm diameter sphere, and V^Emph0^ in this volume that must be 10% or more smaller than V^Emph5^ [=100*(V^Emph0^ - V^Emph5^)/V^Emph5^]. These constraints indicate that these lung regions had sufficient emphysema and progression over the 5-year period; 3. The V^Emph5^ segmentation map was spatially aligned to the baseline expiratory CT scan using LDA software; 4. The aligned V^Emph5^ segmentation map was multiplied to the baseline lobe segmentation map to generate a lobe-specific emphysema map. Detailed inclusion criteria for subjects are provided in **Supplemental Figure 1**.

### Data and Statistical Analysis

Data are presented as mean and standard deviations unless stated otherwise. Statistical work was undertaken using MATLAB R2019a and IBM SPSS Statistics v27 (SPSS Software Products). In all tests, significance was defined by p < 0.05.

### Lobar Cluster Analysis of Emphysema Regions

We assumed that the topology of PRM^Norm^ and PRM^fSAD^ at baseline provided sufficient information to represent all PRM classifications. As such, eight features (i.e., V_i_, S_i_, B_i_, and χ_i_, where i represents PRM^Norm^ and PRM^fSAD^ at baseline) were included in an unsupervised cluster analysis. This analysis was performed using a K-means algorithm. Individual lobes with emphysema involvement were treated as independent measures. The number of clusters was objectively determined using the Calinski-Harabasz method. The relative contributions of each cluster by lobe were determined and evaluated. Cluster differences in baseline tPRM measures, lesion volume, V^Emph^ at year 5 and change in V^Emph^ normalized to V^Emph5^ were determined using Kruskal-Wallis test.

### Subject Subtype Analyses

As cluster analysis was performed at the lobe-level, an individual case may consist of all three clusters. Subjects were designated into three groups based on the following criteria: subjects with lobe-level cluster 2 (2, 12, 23, and 123) were designated emphysema-dominant (ED), remaining subclusters with lobe-level cluster 1 (1 and 13) were designated fSAD-dominant (FD), and the rest (3) were designated fSAD-transition (FT) (see **Supplemental Figure 4**). Differences in various continuous and categorical variables between subject groups were determined using the Kruskal-Wallis non-parametric test with Bonferroni post-hoc testing and Pearson χ_2_ test, respectively.

### Institutional Review Board Approval Statement

Our study was a secondary analysis of data from COPDGene (ClinicalTrials.gov: NCT00608764), a large NIH-funded prospective multi-center observational study. In Phase 1 (2007-2012) of the original study, written and informed consent was obtained from all participants and the study was approved by local institutional review boards of all 21 centers.

## RESULTS

**Table 1** provides total subject characteristics at the time of Phase 1 accrual. This population (N=1495) was predominantly male with a mean age and BMI, with standard deviation (SD), of 62 ± 8 years and 27 ± 5 kg/m^2^, respectively. Most subjects had mild to moderate COPD (GOLD 1 and 2), with only 4% of the population diagnosed with very severe COPD (GOLD 4). At-risk subjects with > 10 pack year smoking history and no COPD diagnosis made up a quarter of the cohort (N = 367). PRM classifications in the entire cohort were primarily normal (49 ± 16%) and fSAD (22 ± 12%).

**Table 1:**
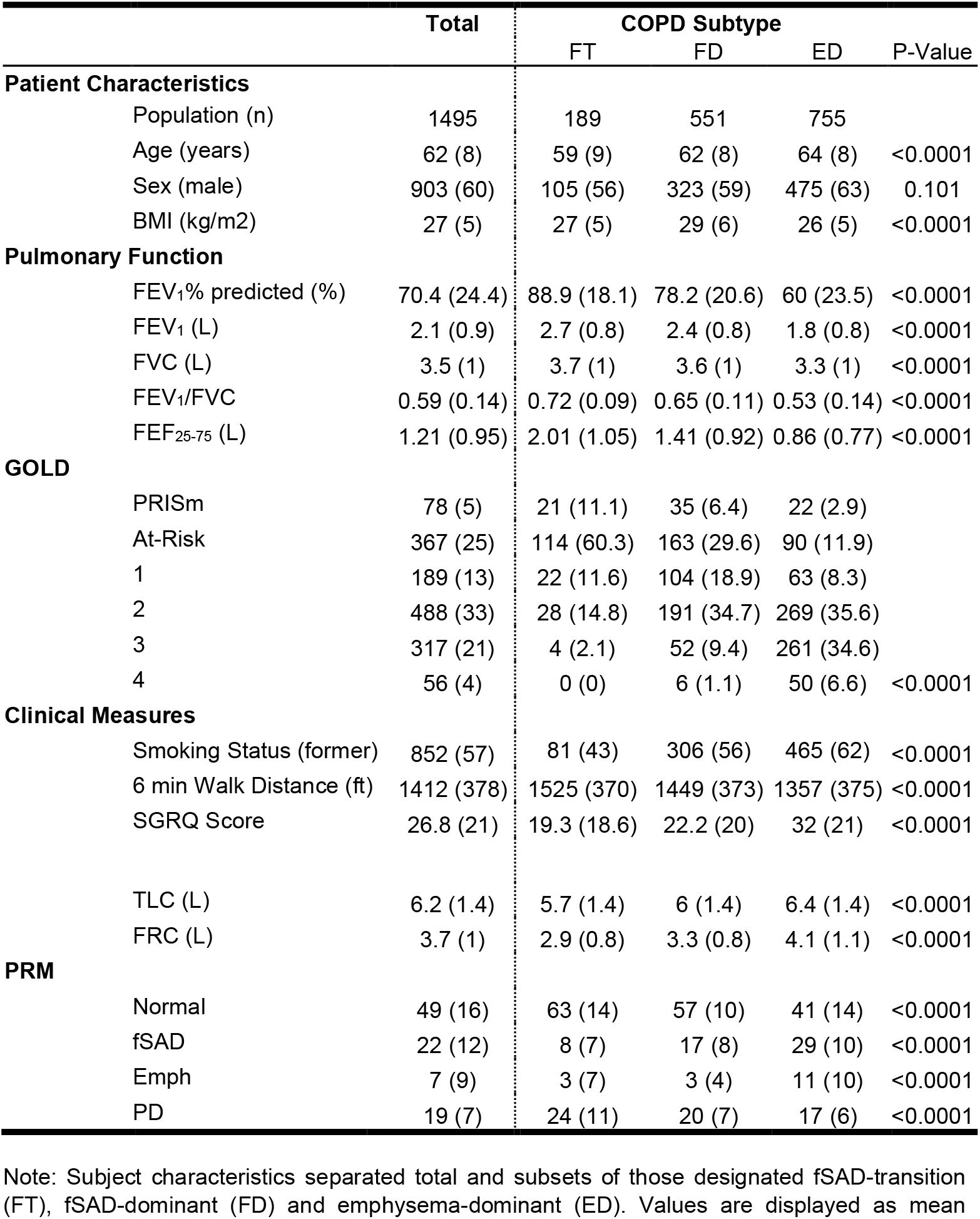
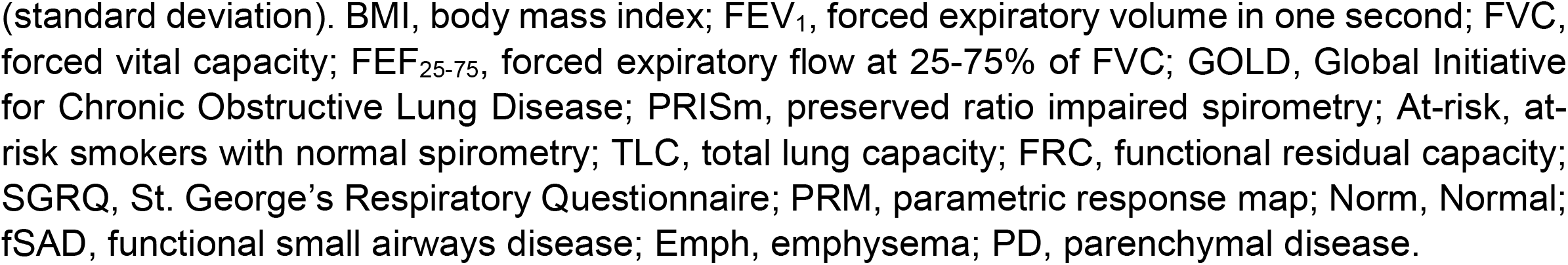
Subject Characteristics

### Lobar-Based Cluster Analysis of CT Regions

As emphysema progression is a local event, each lobe with emphysema was treated as an independent observation. As such, a sole case may have up to 5 observations, representing emphysema involvement in all 5 lobes. We identified 3 well-defined clusters using only topology readouts from PRM^Norm^ and PRM^fSAD^ representing distinct imaging patterns. Additional details are provided on cluster methods and results in the **Supplemental Results**.

Presented in **Table 2** are the lobe-specific characteristics for each imaging cluster. It is important to note that the mean volume density is proportional to the percentage of PRM for a given volume (%PRM^i^ = 100*V^i^, where i indicates a PRM class). Sorted by total observations, clusters 1 and 2 had a similar number of observations, with roughly a third observed for cluster 3 (**Table 2**). Cluster 2 was found to have the largest emphysema volume (0.13 ± 0.129 L) and volume density (V^Emph^) at year 5 (0.43 ± 0.1). Emphysema volume, volume density and change were found to be significantly different between all clusters (pair-wise p<0.0001). Cluster 3 demonstrated the largest percentage difference in V^Emph^ at baseline normalized to year 5 [defined as 100*(V^Emph(yr0)^ – V^Emph(yr5)^)/V^Emph(yr5)^]. No noticeable lobe preference for clusters was observed.

**Table 2:** Lobar-Based Cluster Analysis of CT Regions to Identify Unique Imaging Patterns

|  | Clusters |  |  | P-Values |
| --- | --- | --- | --- | --- |
|  | 1 | 2 | 3 |  |
| **Total (N)** | 1761 | 1711 | 631 |  |
| **Emphysema** |  |  |  |  |
| Volume (L) at year 5 | 0.05 (0.07) | 0.13 (0.13) | 0.02 (0.05) | <0.0001 |
| $V^{\text{Emph}}$ at year 5 | 0.35 (0.06) | 0.43 (0.1) | 0.36 (0.08) | <0.0001 |
| $\Delta V^{\text{Emph}}$ relative to $V^{\text{Emph}}$ at 5 (%) | -49.9 (21.2) | -33.1 (17.8) | -57.5 (27) | <0.0001 |
| **Percentage of Lobe Observations per Cluster (%)** |  |  |  |  |
| RUL | 18 | 21 | 22 |  |
| RLL | 21 | 16 | 22 |  |
| RML | 20 | 24 | 14 |  |
| LUL | 21 | 23 | 23 |  |
| LLL | 19 | 16 | 19 |  |

Note: Lobar-based cluster results are presented as counts or means (SD). Each lobe was considered an independent observation such that each individual may have up to 5 observations. Continuous variables include $V^{\text{Emph}}$ , volume density of PRM<sup>Emph</sup>; $\Delta V^{\text{Emph}}$ relative to $V^{\text{Emph}}$ at 5 (%), difference of $V^{\text{Emph}}$ from baseline to year 5 normalized to year 5 values [ $100 \times (V^{\text{Emph}}_{\text{baseline}} - V^{\text{Emph}}_{\text{at year 5}}) / V^{\text{Emph}}_{\text{at year 5}}$ ]; and Percentage of Lobe Observations per Cluster, the sum of a cluster in a lobe normalized to the sum of the same cluster in all lobes\*100. RUL, right upper lung; RLL, right lower lung; RML, right middle lung; LUL, left upper lung; and LLL, left lower lung.

**Figure 1** shows the cluster results of topology readouts volume density (V) and Euler-Poincaré Characteristic (χ) for all PRM classifications. In brief, V quantifies the amount of a PRM classification, whereas χ quantifies the consolidation of a PRM classification into small pockets (positive values) or a large mesh (negative values). As seen in **Figure 1A**, cluster 2 showed the highest V^Emph^ (0.29 ± 0.11), which was accompanied by the highest levels of V^fSAD^ (0.43 ± 0.11). Lobe Cluster 1 consisted of high levels of V^fSAD^ (0.35 ± 0.09) and cluster 3 consisted mostly of V^Norm^ (0.41 ± 0.16). In **Figure 1B**, Lobe Cluster 2 had the largest negative value in χ^fSAD^ (−0.008 ± 0.008). In contrast, elevated levels in χ, associated with formation of pockets, were only observed for χ^fSAD^ (0.010 ± 0.007) in cluster 3 and χ^Emph^ (0.012 ± 0.007) in Lobe Cluster 1. Surface area (S) and mean breadth (B) also showed unique combinations in values for PRM-derived Norm and fSAD (**Supplemental Figure 3**). All topologies for PRM^Norm^ and PRM^fSAD^ were statistically different between clusters (p<0.0001).

**Figure 1:**
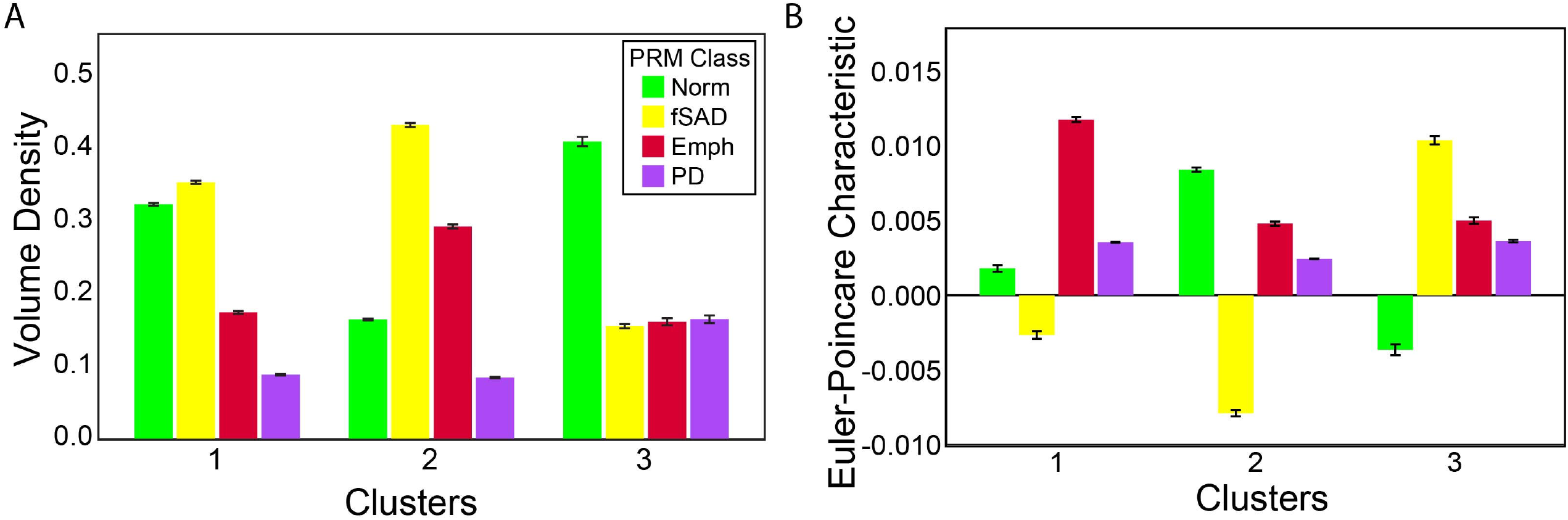
Bar plots for (A) Volume Density (V) and (B) Euler-Poincaré Characteristic (χ) of all PRM classifications across all clusters. Data are presented as mean and SD. PRM classifications include Norm, normal lung parenchyma (green); fSAD, functional small airways disease (yellow); Emph, emphysema (red); and PD, parenchymal disease (magenta).

**Figure 2:**
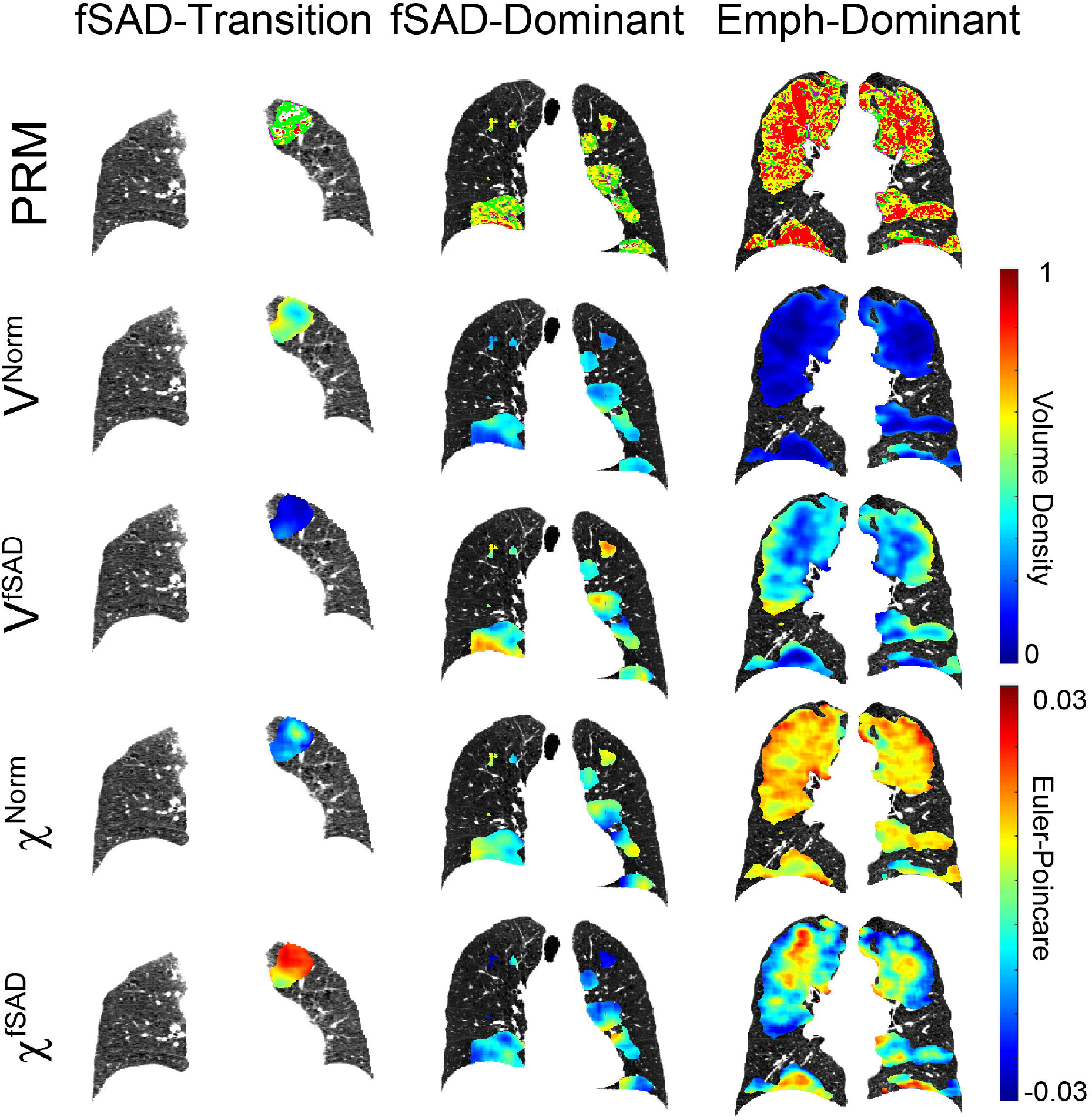
Volume Density (V) and Euler-Poincaré Characteristic (χ) for PRM^Norm^ and PRM^fSAD^ in all determined subtypes. For each subtype, representative coronal slices are provided for the aligned inspiration CT scan acquired at baseline with overlays of PRM, V^Norm^, V^fSAD^, χ^Norm^ and χ^fSAD^. The fSAD-transition (FT) case is a female, 51-55 years of age at enrollment with FEV_1_% predicted of 105% identified as at-risk (i.e., GOLD 0). The fSAD-dominant (FD) case is a female (51-55 years old) with FEV_1_% predicted of 84% diagnosed with GOLD 1 COPD. The emphysema-dominant (ED) case is a male (56-60 years old) with FEV_1_% predicted of 31% diagnosed with GOLD 3.

**Figure 3:**
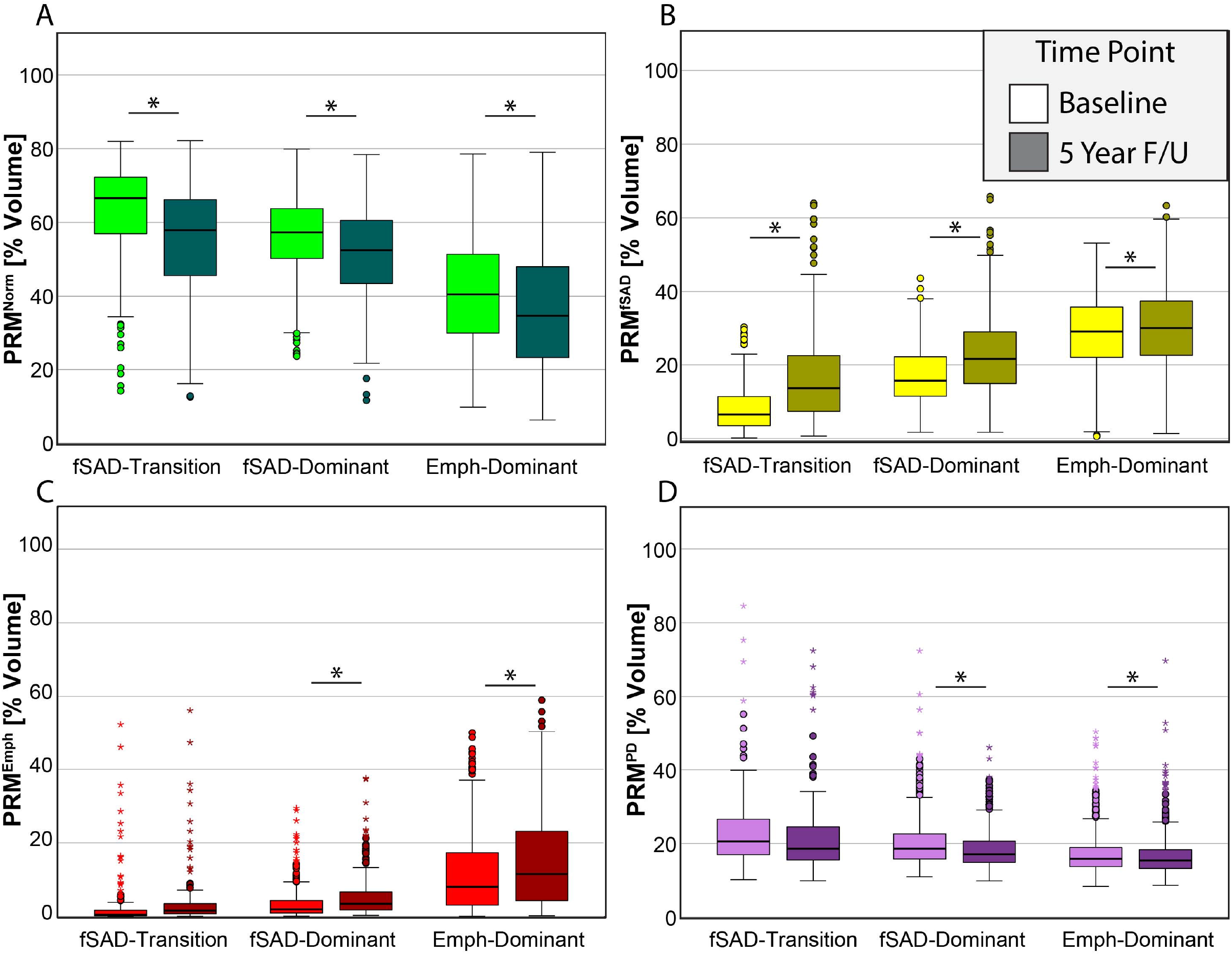
Box and whisker plots for Phase 1 and 2 measurements of the percent volume of PRM classifications (A) Norm (normal parenchyma; color coded green), (B) fSAD (functional small airways disease, color coded yellow), (C) emphysema (color coded red) and (D) PD (parenchymal disease; color coded magenta). Box represents the 25^th^ and 75^th^ percentiles, line represents median, and whiskers represent minimum and maximum values. Circles and stars represent outliers and extreme values. * indicates significant difference between time intervals at p<0.05.

### Subject Subtype Analyses

We observed varying contributions of fSAD and emphysema in our imaging clusters. Lobe Cluster 2, which we identified as emphysema dominant, exhibited elevated V^Emph^ and V^fSAD^. Lobe Cluster 1, fSAD dominant, had high V^fSAD^, low V^Emph^ and elevated χ^Emph^ (emphysema pockets). Finally, Lobe Cluster 3, transition of normal parenchyma to fSAD, consisted of low V^Emph^ and V^fSAD^ but elevated χ^fSAD^. Depending on lobe cluster involvement, we grouped individual subjects into subtypes as follows: fSAD-transition (FT), fSAD-dominant (FD), and emphysema-dominant (ED), (details are provided in **Supplemental Figure 4**). **Figure 2** highlights three representative subjects, each from a different subtype. The subject designated as ED was diagnosed with GOLD 3 COPD (FEV_1_% predicted of 31%) and had whole-lung percent volumes of PRM^Emph^ and PRM^fSAD^ of 26% and 41%, respectively. The FD subject was diagnosed with GOLD 1 COPD (FEV_1_% predicted of 84%) with percent volumes of PRM^Emph^ and PRM^fSAD^ of 2% and 32%, respectively. The subject designated as the FT subtype had a low symptom burden as measured by SGRQ score (GOLD 0, FEV_1_% predicted and FEV_1_/FVC of 105% and 0.75, respectively) with negligible PRM^Emph^ (0.6%) and PRM^fSAD^ (1%). At Phase 2, the FT subject progressed to GOLD 1 COPD (FEV_1_/FVC of 0.66).

Subject characteristics at baseline are presented in **Table 1** by subtypes. All variables except sex were found to be significantly different between subtypes. With respect to pulmonary function measurements, ED had the lowest values in all measurements. FD had lower pulmonary function measurements than FT. However, subjects from every GOLD stage were seen in every subtype, suggesting these patient designations do not simply represent differences in disease severity. ED did show the highest prevalence of GOLD 3 and 4 subjects (41.2%), which accounted for elevated whole-lung PRM^Emph^ (11 ± 10%). FD showed nearly three times as many at-risk subjects (29.6%) with nearly as many GOLD 2 (34.7%) as ED (11.9% and 35.6%, respectively). In contrast, FT consisted predominantly of at-risk subjects, which made up 60.3% compared to 29.6% and 11.9% for FD and ED, respectively. Six-minute walk and SGRQ scores differed significantly between subsets except for FD and FT (p=0.056 and 0.215, respectively). As expected, whole-lung PRM values aligned with our subject subset designations. All PRM and CT lung volumes were significantly different between subtypes.

We further evaluated changes in PRM and pulmonary function testing (PFT) measures over the 5-year period within subjects identified in each subtype. Subjects designated as having FD and ED were found to demonstrate significant changes in all four PRM classifications (p<0.05; **Figure 3**). Subjects in the FT subtype were found to increase and decrease significantly only in PRM^Norm^ (**Figure 3A**) and PRM^fSAD^ (**Figure 3B**), respectively. Evaluating the percent change in PRM classifications between subtypes (**Table 3**) showed that subjects designated as ED demonstrated an increase in PRM^Emph^ of 3.8%, which was significantly higher than the other subtypes. The largest change in fSAD was observed in FT (9.0 +/-13.7%), followed by FD (5.7 +-8.9%), which were statistically similar (p=0.536), but significantly larger than ED (p<0.0001 for both). No significant differences were observed between subtypes for changes in FEV_1_. Those cases designated as ED demonstrated the highest rates of change in FVC but were only found to be significant with FT (p=0.011). FD and FT subtypes showed rates of FEF_25-75_ (forced expiratory flow at 25-75% of FVC) decline that were significantly higher than ED (p=0.006 and p<0.0001, respectively), but these rates were not found to differ significantly (p=0.104).

**Table 3:** Change in Whole-Lung PRM and Pulmonary Function Measurements

|  | Subtypes |  |  | P-Value |
| --- | --- | --- | --- | --- |
|  | FT | FD | ED |  |
| Parametric Response Map [%] |  |  |  |  |
| Norm | -8.4 (14.6) | -5.2 (10) | -4.3 (8.3) | 0.027 |
| fSAD | 9 (13.7) | 5.7 (8.9) | 1.4 (7.8) | <0.0001 |
| Emph | 1.2 (2.4) | 1.8 (3.2) | 3.8 (4.9) | <0.0001 |
| PD | -1.6 (7.8) | -1.9 (5.1) | -0.7 (3.7) | <0.0001 |
| Pulmonary Function [mL/yr] |  |  |  |  |
| FEV<sub>1</sub> | -45.2 (61.2) | -50.7 (60.6) | -47.7 (58.5) | 0.101 |
| FVC | -45.6 (84.7) | -62.1 (85.1) | -67.9 (104) | 0.014 |
| FEF<sub>25-75</sub> | -54.9 (116.5) | -36.1 (98.1) | -23.3 (68.1) | <0.0001 |

Note: Change in whole-lung PRM and pulmonary function measurements separated by subsets of those designated fSAD-transition (FT), fSAD-dominant (FD) and emphysema-dominant (ED). Values are displayed as mean (standard deviation). Norm, Normal; fSAD, functional small airways disease; Emph, emphysema; PD, parenchymal disease; FEV<sub>1</sub>, forced expiratory volume in one second; FVC, forced vital capacity; FEF<sub>25-75</sub>, forced expiratory flow at 25-75% of FVC.

## DISCUSSION

COPD is characterized by significant heterogeneity in the amount of airway disease and emphysema. PRM can distinguish between fSAD and emphysema, and as we demonstrate in this work, local emphysema progression can be identified by assessing the topology (i.e., amount and arrangement) of PRM-defined normal parenchyma (PRM^Norm^) and fSAD (PRM^fSAD^) [21] to better understand how and in whom COPD progresses. The focus on both lobar level and subject specific analyses reveals important overarching themes. Broadly, we have identified 3 unique radiologic patterns: fSAD-dominant (FD), emphysema-dominant (ED), and fSAD-transition (FT)— the latter a group consisting of a unique transitional state from normal lung to fSAD that is associated with emphysema progression.

Emphysema-dominant subjects had the lowest baseline pulmonary function measurements, an expected result given that these subjects had the highest degree of emphysema at study enrollment. This subtype also had the highest prevalence of severe COPD subjects with the greatest number of GOLD 3 and 4 subjects. Not surprisingly, ED subjects were more likely to have ever smoked and had the lowest functional class (with the lowest 6-minute walk distance) and highest degree of symptom burden (with the highest SGRQ score). These were all statistically significant differences among the three subtypes. ED subjects had the largest increase in whole-lung PRM-defined emphysema after 5 years, which was also statistically significantly different (**Table 3**). While many of these findings are expected in this group with advanced disease, it was interesting to note that these radiologic changes correlated to worsening symptoms and functional status, but not necessarily to large changes in lung function after accounting for aging. This lack of significance further emphasizes that spirometric changes cannot consistently account for the degree of radiologic changes identified by PRM in subjects at risk for or with COPD over a short follow up period [23]. These findings suggest that tPRM may provide earlier evidence of regional disease progression and reveal different clinical trajectories, and thus, represent a more sensitive tool than global PFTs alone.

Lobes identified as fSAD-dominant showed nearly a 50% regional change in emphysema in the 5-year follow up period. This was greater than what was seen in the emphysema-dominant cluster 2, which only had a 33% increase in emphysema (**Table 2**). This is consistent with recent studies showing that in cigarette smokers without baseline emphysema, the presence of fSAD is associated with emphysema development [24]. Over half of the subjects in the FD group were GOLD 1 or 2; however, 30% of subjects in this group were at-risk subjects, or GOLD 0. This suggests that a significant number of subjects at risk for developing COPD and with mild-moderate COPD all have some degree of fSAD. χ^Emph^ was high in the FD group, indicating the presence of small pockets of emphysema developing within larger regions of fSAD. As these subjects may not have yet progressed to end stage disease with a large amount of irreversible emphysema, they may fall into a separate group where novel therapies and close monitoring may further improve their quality of life and clinical outcomes. FD subjects also had the greatest change in FEV_1_ at nearly a 50 mL/year drop; however, this was not significantly different compared to the other groups. This indicates PFT metrics may not be able to capture overall global functional changes in a short interval, as emphysema develops locally from regions of fSAD and these regions of fSAD also evolve from healthy lung tissue.

The discovery with the greatest potential clinical utility is arguably the FT subtype, cluster 3. These clusters had even larger regional increases in emphysema over this 5-year follow up period—nearly 58%**–**the largest among all the groups (see **Supplemental Results** for cluster analysis). The FT subtype comprised of subjects with predominantly little emphysema or fSAD as measured using volume density. In contrast, χ^fSAD^ was found to be elevated, suggesting the presence of fSAD pockets already present within healthy lung parenchyma at study enrollment [25]. Many prior studies have shown that GOLD 0 subjects consistently have both clinical and radiologic evidence of smoking-related disease [26]; however, it remains difficult to identify these changes with currently available tools. FT subjects’ pulmonary function testing revealed that their FEV_1_/FVC ratio and FEV_1_% predicted were largely preserved at baseline, again consistent with the at-risk label. FEF_25-75_% decline in the FT subtype was also the greatest of all the subtypes, supporting the theory that it may be an early marker of COPD development [25, 27]. These at-risk subjects still have a significant symptom burden despite normal spirometry, which may be reflected in the pockets of abnormal airway remodeling we visualized in this study.

This large-scale study was the first to use tPRM across Phases 1 and 2 of COPDGene, which was comprised of a cohort of diverse subjects across the country. One of the strengths of this work is our strategy to evaluate tPRM readouts in lung regions with confirmed emphysema progression over 5 years. This allowed us to identify new emphysema phenotypes, particularly our description of a unique transitional stage between healthy lung and development of fSAD (i.e., FT), which has not been previously discussed in the literature. Furthermore, we have shown that these different clinical phenotypes both correlate with and add insight to available PFT data that may allow clinicians to phenotype patients earlier in their disease courses. One of the intriguing possibilities of tPRM is the ability to quantify regional risk of emphysema progression over time in a way that global PFT metrics cannot do. Even in subjects with advanced COPD, resulting in severe obstruction and gas exchange impairment, there may be regions in the lung with reversible damage (such as in FT clusters with pockets of fSAD) that can be potential therapeutic targets for intervention.

This current study has several limitations. We identified unique disease subtypes based on the topology of PRM classification maps generated from high-resolution CT data from a well-controlled multi-center observational COPD trial. However, different reconstruction kernels and scanner systems are known to result in variations in HU values, which affect the PRM classification maps and resulting topology calculations [28]. In addition, image resolution is critical for topological comparisons, as lower resolution intrinsically appears more clustered, biasing the feature patterns in the CT image. Minimal variation in image resolution was found between data sets for this study. Nevertheless, care was taken to account for image noise and registration errors while assessing our metrics [28]. Despite these limitations, our results have physiologic and clinical correlates that still allow us to draw important conclusions.

There remains more to be explored to extend this work. The FT subtype we described is not yet clinically well-defined and further characterization of whether this phenotype is truly on a spectrum of disease between healthy and development of fSAD and emphysema remains to be confirmed. While many of these FT GOLD 0 subjects may be at increased risk of developing COPD, the link between the pathophysiology of this disease progression as it relates to radiologic changes needs to be further studied. In addition to validating our results with pulmonary function testing, we hope to correlate these findings to other clinically meaningful outcomes, including risk of functional decline, morbidity metrics, COPD exacerbations (especially hospitalizations), and mortality. Studying outcomes beyond the 5-year follow up we look at here will allow us to better understand the longer-term implications of this novel subtype in a disease process that we are increasingly appreciating as heterogenous.

## CONCLUSIONS

Local topological parametric response maps identified three distinctive radiologic tissue patterns that can be used to identify corresponding individual subjects characterized by unique clinical features. This work highlights the discovery of the fSAD transition (FT) subtype, characterized by high χ of fSAD, which may help identify individuals without spirometrically-defined COPD who remain at-risk. Further work is needed to better understand this novel phenotype and its clinical implications in the context of emphysema development and progression. (standard deviation). BMI, body mass index; FEV_1_, forced expiratory volume in one second; FVC, forced vital capacity; FEF_25-75_, forced expiratory flow at 25-75% of FVC; GOLD, Global Initiative for Chronic Obstructive Lung Disease; PRISm, preserved ratio impaired spirometry; At-risk, at-risk smokers with normal spirometry; TLC, total lung capacity; FRC, functional residual capacity; SGRQ, St. George’s Respiratory Questionnaire; PRM, parametric response map; Norm, Normal; fSAD, functional small airways disease; Emph, emphysema; PD, parenchymal disease.

## Data Availability

The datasets presented in this study are not readily available because they are part of NIH sponsored clinical trials and require a data use agreement to be signed. For access to COPDGene data visit https://www.copdgene.org/phase-1-study-documents.htm for instructions.

## Author Contributions

CJG collected data. CJG and JMW designed the analysis and analyzed the data. CJG and JMW drafted the manuscript with editing from MKH. All authors reviewed and approved the final version of the manuscript for publication.

## Funding

This work was supported by the National Heart, Lung, and Blood Institute (NHLBI) of the National Institutes of Health Grants R01HL139690 and R01 HL150023 and by NHLBI Grants U01 HL089897 and U01 HL089856, which support the COPDGene study. The COPDGene study (NCT00608764) is also supported by the COPD Foundation through contributions made to an Industry Advisory Committee comprised of AstraZeneca, Bayer Pharmaceuticals, Boehringer-Ingelheim, Genentech, GlaxoSmithKline, Novartis, Pfizer and Sunovion.

We acknowledge the COPDGene investigators for their role in the study providing data for this project:

## Administrative Center

James D. Crapo, MD (PI); Edwin K. Silverman, MD, PhD (PI); Barry J. Make, MD; Elizabeth A. Regan, MD, PhD

## Genetic Analysis Center

Terri Beaty, PhD; Ferdouse Begum, PhD; Peter J. Castaldi, MD, MSc; Michael Cho, MD; Dawn L. DeMeo, MD, MPH; Adel R. Boueiz, MD; Marilyn G. Foreman, MD, MS; Eitan Halper-Stromberg; Lystra P. Hayden, MD, MMSc; Craig P. Hersh, MD, MPH; Jacqueline Hetmanski, MS, MPH; Brian D. Hobbs, MD; John E. Hokanson, MPH, PhD; Nan Laird, PhD; Christoph Lange, PhD; Sharon M. Lutz, PhD; Merry-Lynn McDonald, PhD; Margaret M. Parker, PhD; Dmitry Prokopenko, Ph.D; Dandi Qiao, PhD; Elizabeth A. Regan, MD, PhD; Phuwanat Sakornsakolpat, MD; Edwin K. Silverman, MD, PhD; Emily S. Wan, MD; Sungho Won, PhD

## Imaging Center

Juan Pablo Centeno; Jean-Paul Charbonnier, PhD; Harvey O. Coxson, PhD; Craig J. Galban, PhD; MeiLan K. Han, MD, MS; Eric A. Hoffman, Stephen Humphries, PhD; Francine L. Jacobson, MD, MPH; Philip F. Judy, PhD; Ella A. Kazerooni, MD; Alex Kluiber; David A. Lynch, MB; Pietro Nardelli, PhD; John D. Newell, Jr., MD; Aleena Notary; Andrea Oh, MD; Elizabeth A. Regan, MD, PhD; James C. Ross, PhD; Raul San Jose Estepar, PhD; Joyce Schroeder, MD; Jered Sieren; Berend C. Stoel, PhD; Juerg Tschirren, PhD; Edwin Van Beek, MD, PhD; Bram van Ginneken, PhD; Eva van Rikxoort, PhD; Gonzalo Vegas Sanchez-Ferrero, PhD; Lucas Veitel; George R. Washko, MD; Carla G. Wilson, MS;

## PFT QA Center, Salt Lake City, UT

Robert Jensen, PhD

## Data Coordinating Center and Biostatistics, National Jewish Health, Denver, CO

Douglas Everett, PhD; Jim Crooks, PhD; Katherine Pratte, PhD; Matt Strand, PhD; Carla G. Wilson, MS

## Epidemiology Core, University of Colorado Anschutz Medical Campus, Aurora, CO

John E. Hokanson, MPH, PhD; Gregory Kinney, MPH, PhD; Sharon M. Lutz, PhD; Kendra A. Young, PhD

## Mortality Adjudication Core

Surya P. Bhatt, MD; Jessica Bon, MD; Alejandro A. Diaz, MD, MPH; MeiLan K. Han, MD, MS; Barry Make, MD; Susan Murray, ScD; Elizabeth Regan, MD; Xavier Soler, MD; Carla G. Wilson, MS

## Biomarker Core

Russell P. Bowler, MD, PhD; Katerina Kechris, PhD; Farnoush Banaei-Kashani, Ph.D

The authors wish to thank Lee Olsen for assisting with manuscript preparation and editing.

## REFERENCES

1. Symposium, A.C.G., Terminology, Definitions, and Classification of Chronic Pulmonarey Emphysema and Related Conditions. Thorax, 1959. 14: p. 286–299.

2. Mohamed Hoesein, F.A., et al., CT-quantified emphysema in male heavy smokers: association with lung function decline. Thorax, 2011. 66(9): p. 782–7.

3. Washko, G.R., et al., Computed tomographic-based quantification of emphysema and correlation to pulmonary function and mechanics. COPD, 2008. 5(3): p. 177–86.

4. Rambod, M., et al., Six-minute walk distance predictors, including CT scan measures, in the COPDGene cohort. Chest, 2012. 141(4): p. 867–75.

5. Diaz, A.A., et al., Relationship of emphysema and airway disease assessed by CT to exercise capacity in COPD. Respir Med, 2010. 104(8): p. 1145–51.

6. Haruna, A., et al., CT scan findings of emphysema predict mortality in COPD. Chest, 2010. 138(3): p. 635–40.

7. Zulueta, J.J., et al., Emphysema scores predict death from COPD and lung cancer. Chest, 2012. 141(5): p. 1216–23.

8. Chabat, F., G.Z. Yang, and D.M. Hansell, Obstructive lung diseases: texture classification for differentiation at CT. Radiology, 2003. 228(3): p. 871–7.

9. Park, Y.S., et al., Texture-based quantification of pulmonary emphysema on highresolution computed tomography: comparison with density-based quantification and correlation with pulmonary function test. Invest Radiol, 2008. 43(6): p. 395–402.

10. Uppaluri, R., et al., Quantification of pulmonary emphysema from lung computed tomography images. Am J Respir Crit Care Med, 1997. 156(1): p. 248–54.

11. Sorensen, L., S.B. Shaker, and M. de Bruijne, Quantitative analysis of pulmonary emphysema using local binary patterns. IEEE Trans Med Imaging, 2010. 29(2): p. 559–69.

12. Castaldi, P.J., et al., Distinct quantitative computed tomography emphysema patterns are associated with physiology and function in smokers. Am J Respir Crit Care Med, 2013. 188(9): p. 1083–90.

13. McDonough, J.E., et al., Small-Airway Obstruction and Emphysema in Chronic Obstructive Pulmonary Disease. New England Journal of Medicine, 2011. 365(17): p. 1567–1575.

14. Galban, C.J., et al., Computed tomography-based biomarker provides unique signature for diagnosis of COPD phenotypes and disease progression. Nat Med, 2012. 18(11): p. 1711–5.

15. Labaki, W.W., et al., Voxel-Wise Longitudinal Parametric Response Mapping Analysis of Chest Computed Tomography in Smokers. Acad Radiol, 2019. 26(2): p. 217–223.

16. Regan, E.A., et al., Genetic Epidemiology of COPD (COPDGene) Study Design. COPD: Journal of Chronic Obstructive Pulmonary Disease, 2011. 7(1): p. 32–43.

17. Rabe, K.F., et al., Global Strategy for the Diagnosis, Management, and Prevention of Chronic Obstructive Pulmonary Disease. American Journal of Respiratory and Critical Care Medicine, 2007. 176(6): p. 532–555.

18. Wan, E.S., et al., Epidemiology, genetics, and subtyping of preserved ratio impaired spirometry (PRISm) in COPDGene. Respir Res, 2014. 15: p. 89.

19. Jones, P.W., et al., A self-complete measure of health status for chronic airflow limitation. The St. George’s Respiratory Questionnaire. Am Rev Respir Dis, 1992. 145(6): p. 1321–7.

20. Belloli, E.A., et al., Parametric Response Mapping as an Imaging Biomarker in Lung Transplant Recipients. American Journal of Respiratory and Critical Care Medicine, 2016. 195(7): p. 942–952.

21. Hoff, B.A., et al., CT-Based Local Distribution Metric Improves Characterization of COPD. Scientific Reports, 2017. 7(1): p. 2999.

22. Legland, D., K. Kiêu, and M.-F. Devaux, COMPUTATION OF MINKOWSKI MEASURES ON 2D AND 3D BINARY IMAGES. Image Analysis & Stereology, 2007. 26(2): p. 83–92.

23. Pompe, E., et al., Five-year Progression of Emphysema and Air Trapping at CT in Smokers with and Those without Chronic Obstructive Pulmonary Disease: Results from the COPDGene Study. Radiology, 2020. 295(1): p. 218–226.

24. Pompe, E., et al., Progression of Emphysema and Small Airways Disease in Cigarette Smokers. Chronic Obstr Pulm Dis, 2021. 8(2): p. 198–212.

25. Han, M.K., et al., From GOLD 0 to Pre-COPD. Am J Respir Crit Care Med, 2021. 203(4): p. 414–423.

26. Regan, E.A., et al., Clinical and Radiologic Disease in Smokers With Normal Spirometry. JAMA Internal Medicine, 2015. 175(9): p. 1539–1549.

27. Kwon, D.S., et al., FEF25-75% Values in Patients with Normal Lung Function Can Predict the Development of Chronic Obstructive Pulmonary Disease. Int J Chron Obstruct Pulmon Dis, 2020. 15: p. 2913–2921.

28. Boes, J.L., et al., The Impact of Sources of Variability on Parametric Response Mapping of Lung CT Scans. Tomography, 2015. 1(1): p. 69–77.

