## Supplemental Material for "Topologic Parametric Response Mapping Identifies Tissue Subtypes Associated with Emphysema Progression"

**Supplemental Information Guide**

| **Item** | **Title/Description** |
|---|---|
| Supplemental Figure 1 | Study Schema |
| Supplemental Figure 2 | Cluster Analysis |
| Supplemental Figure 3 | Bar Plot for Lobe-Analysis of Surface Area and Mean Curvature |
| Supplemental Figure 4 | Designation of Cases by Lobe-based Clusters |

### Supplemental Figure 1

Study Schema

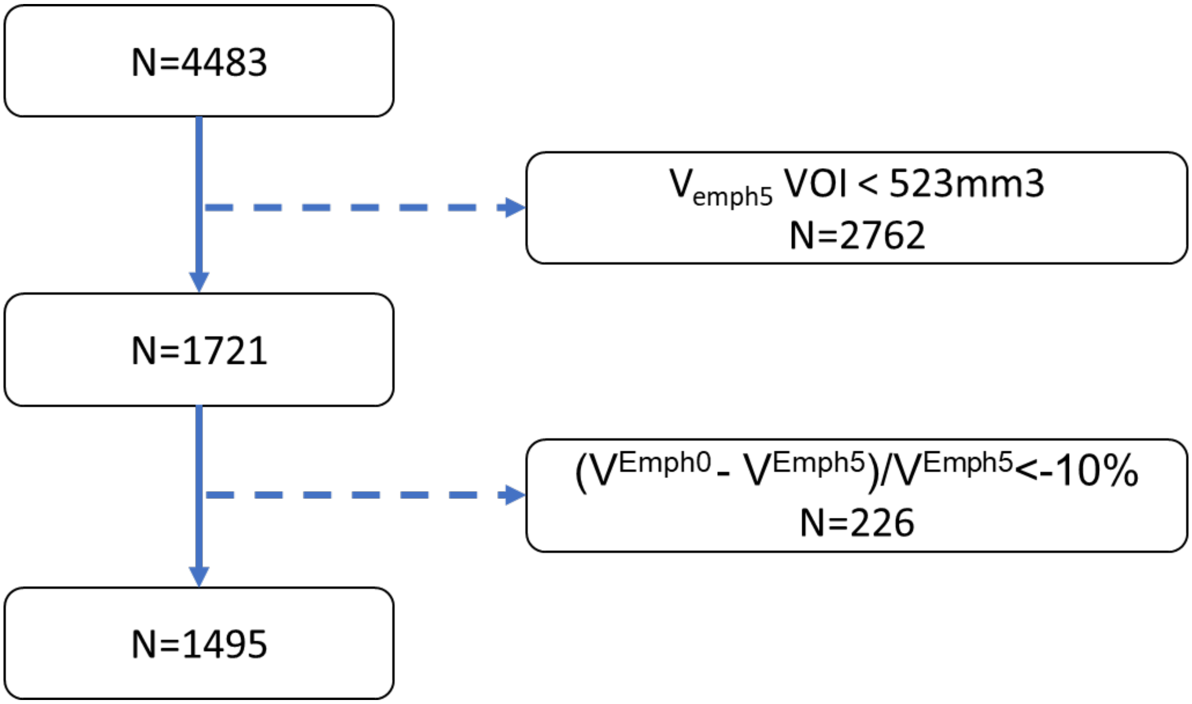


Study schema provides details on attrition of the study population. The criteria for inclusion are: 1. Emphysema volumes, defined by the volume density of PRM-derived emphysema at year 5 ($V_{Emph5}$), must have a volume greater than 523 $mm^3$. This is based on a spherical volume with a diameter of 10 mm; 2. The mean volume density of PRM-defined emphysema at year 0 ($V_{Emph0}$) must be less than 90% of $V_{Emph5}$.

### Supplemental Figure 2

Cluster Analysis

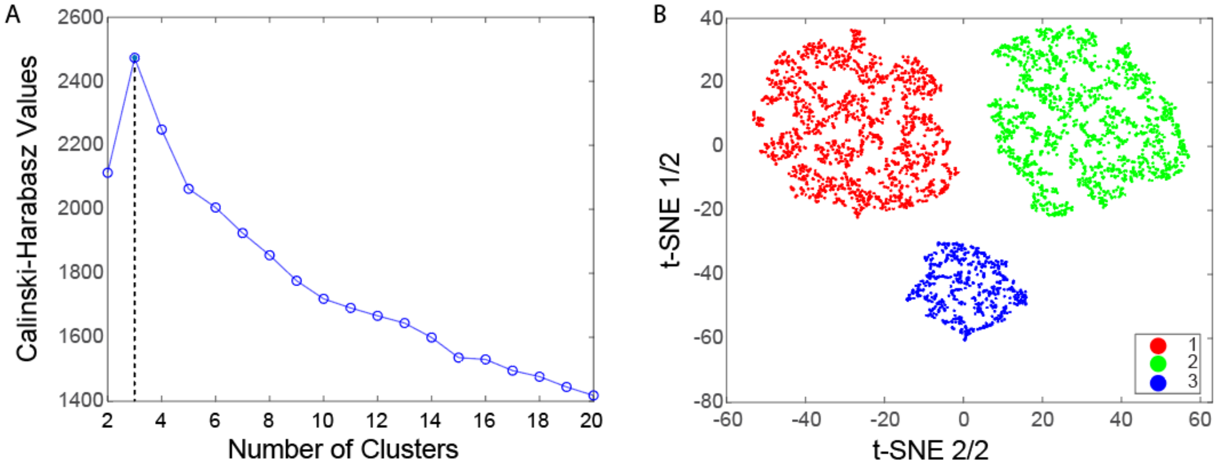


A cluster analysis was performed using a K-means algorithm on the topology readouts (volume density (V), surface area, (S) mean curvature (B) and Euler-Poincaré Characteristics (χ)) for PRM-derived normal and fSAD (8 measurements). The optimal number of clusters was determined objectively using the **(A)** Calinski-Harabasz criterion. To visualize the individual clusters **(B)**, a tSNE algorithm was applied to the 8 readouts including the cluster assignments with results projected onto a 2D space. It is important to note that clustering was not done on the tSNE data itself, which is commonly used for data reduction. It was instead used as a visualization tool, as it projects points from high dimensional space onto (in this case) 2D, so that neighboring points reflect their similarity[1]. All analyses were performed using MATLAB R2019a (MATLAB, The MathWorks Inc., Natick, MA).

#### Supplemental Figure 3

Bar Plot for Lobe-Analysis of Surface Area and Mean Curvature

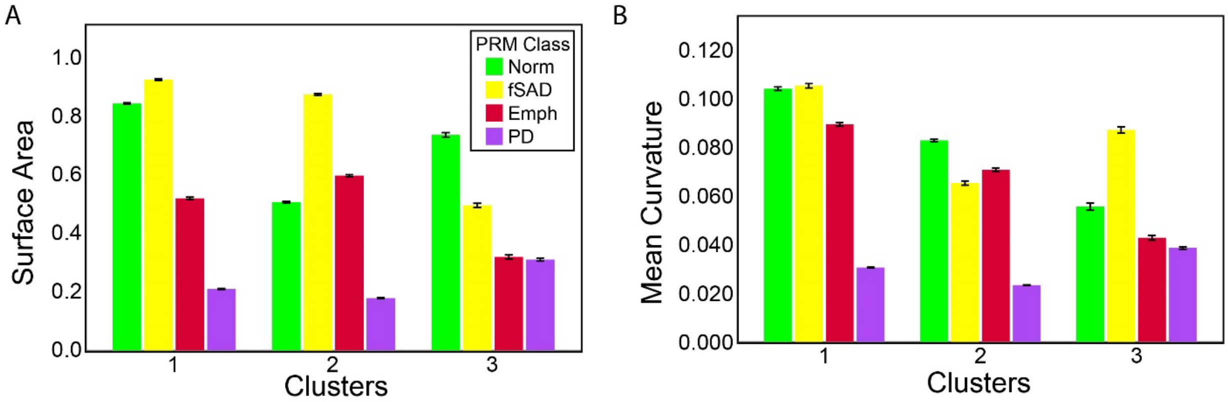


Boxplots for topological measures of PRM maps across all lobe-based clusters. **(A)** surface area, describing class exposure (exposed faces of voxels). **(B)** mean curvature, which can be interpreted as average curvature (convex/concave) of class surfaces. Data presented as means and standard error of the means. PRM classifications are presented as Norm for normal parenchyma (green), fSAD for functional small airways disease (yellow), Emph for emphysema (red) and PD for parenchymal disease (magenta).

### Supplemental Figure 4

Designation of Cases by Lobe-based Clusters

A

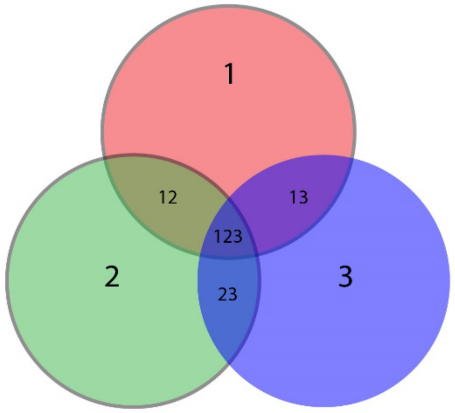


B

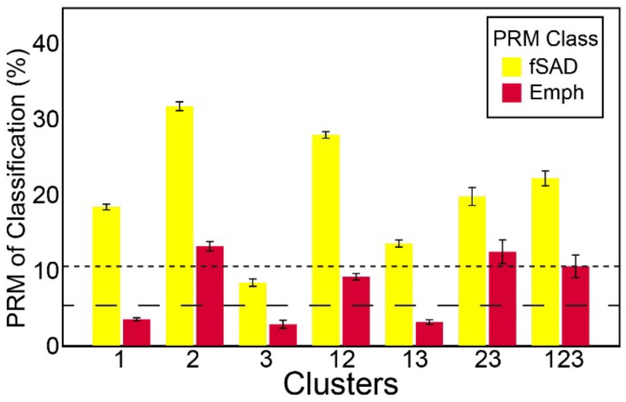


C

| | FT | FD | | ED | | | |
|---|---|---|---|---|---|---|---|
| N | 189 | 421 | 130 | 355 | 296 | 58 | 46 |
| Cluster | 3 | 1 | 13 | 2 | 12 | 23 | 123 |

As cluster analysis was performed at the lobe-level, an individual case may consist of all three clusters. **(A)** The Venn diagram shows the overlap between all three clusters (1, 2, 3), which resulted in 7 possible combinations. **(B)** Bar plot shows the whole-lung levels of PRM-derived emphysema and fSAD for each of the 7 cluster combinations. Large and small dashed lines indicate 5% and 10% of the whole-lung volume. **(C)** Subjects were designated into three groups based on the following criteria: subjects with lobe-level cluster 2 (2, 12, 23, and 123) were designated emphysema-dominant (ED), remaining subclusters with lobe-level cluster 1 (1 and 13) were designated fSAD-dominant (FD), and the rest (3) were designated fSAD-transition (FT).

#### References

1. Balsor JL, Arbabi K, Singh D, et al. A Practical Guide to Sparse k-Means Clustering for Studying Molecular Development of the Human Brain. *Front Neurosci*. 2021;15:668293. doi:10.3389/fnins.2021.668293
